# Implementing Reproductive Carrier Screening to Include Diverse Asian Populations: Insights from Singapore

**DOI:** 10.64898/2026.04.07.26350306

**Authors:** Yasmin Bylstra, Myrabeth Yeo, Jing Xian Teo, Jeannette Goh, Christina Choi, Shermaine Chan, Christine Song, Jasmine Chew Yin Goh, Nellie Bin Siew Chai, Jane Andrea Leviant, Hui Jin Toh, Sock Hoi Chan, Robin Blythe, Melody Menezes, Chengxi Yang, Jan Hodgson, Nicholas Graves, Joel Sng, Wan Wan Lim, Hai Yang Law, David J. Amor, Gareth Baynam, Jerry KY Chan, Yoke Hwee Chan, Patrick Tan, Ivy Ng, Weng Khong Lim, Saumya S. Jamuar

## Abstract

**Background:** As part of Singapore’s effort towards precision medicine tailored to Asian diversity, we describe the implementation of a nationwide reproductive carrier screening program. Using a customised 112-gene panel, incorporating population-specific recessive genetic diseases, we outline the overall program design, and initial efforts of community and stakeholder engagement, to inform culturally appropriate implementation.

**Methods:** Participants receive culturally tailored online education regarding our reproductive screening program and are provided results with genetic counselling and reproductive options. Community and stakeholder perspectives were assessed through questionnaires and consultations with religious leaders.

**Results:** Recruitment is nation-wide, and since initiation of our pilot phase in September 2024, 1,619 couples have registered interest, with 60% uptake of those deemed eligible. Among the 456 couples that have received results to date, four couples (0.9%) were identified to be at increased risk. Community questionnaire responses (n=1002), involving couples who participated in the program as well as the general public, indicated interest is high (59%) across the cohort but awareness, intent to participate and implications for reproductive options differed by sociodemographic factors such as ancestry and religion. Healthcare professional respondents (n=113) acknowledged carrier screening will be routine in medical care, but report limited confidence and resources. Engagement with religious leaders indicated support for the program.

**Conclusion:** These early program outcomes and community engagement are guiding the implementation of expanding population-based carrier screening in Singapore, contingent on addressing practical challenges through equitable outreach and professional training.

## Background

Singapore launched Phase III of its Precision Medicine Programme on 14 November 2025, to expand the implementation of precision medicine for the population(1). This phase will accelerate the adoption of precision medicine in routine clinics, supporting a shift from reactive treatment to proactive and preventative healthcare. Within this framework, population based genomic screening will be integrated to enable personal risk stratification and targeted interventions prior to disease onset. Reproductive carrier screening, and more recently newborn genomic screening(2, 3), are the proverbial low hanging fruit when implementing genomic based preventative healthcare. Success of carrier screening programs such as the Dor Yeshorim in New York showcase the opportunity to reduce the incidence of severe and untreatable yet devastating neurogenetic diseases in the Ashkenazi Jewish community(4). Similarly, the successful implementation of guidelines from professional societies with targeted carrier screening for thalassemia in the Mediterranean, African and Southeast Asian populations, and cystic fibrosis in the non-Jewish Caucasian and Ashkenazi Jewish population(5–7), combined with improved ability to analyse genomic information has led to promulgation of expanded carrier screening, moving beyond ethnicity-based to all recessive genetic diseases(8). However, due to the increased cost of testing all recessive genetic diseases, the adoption of this test as a national screening program is often limited. For example, in Australia, the success of Mackenzie’s Mission expanded screening(9) is available as a charged service however government funding is available for three diseases, namely cystic fibrosis, spinal muscular atrophy and fragile X syndrome(10).

Recent work in Asian populations have identified a different pattern of epidemiology for these recessive diseases. In 2018, we identified citrin deficiency, caused by variants in *SLC25A13*, to be the most common treatable inherited disorder among Southeast Asians, specifically among Singaporeans of Chinese ethnicity(11). A follow up study that analysed 9,051 healthy Singaporeans identified that 27% of genes associated with conditions classified as Tier 3 and above of the American College of Medical Genetics and Genomics (ACMG) guidelines(8) yet were not included in any commercially available carrier screening panels(12).

Based on these findings, we developed a customised carrier screening panel comprising 112 genes across 80 severe recessive genetic disease and were prevalent in the local population(13) and have embarked on a program to implement this panel across 39,000 couples. Through this program, we will study the implementation of carrier screening in an Asian context addressing technical, logistical, ethical and social challenges. This paper describes the design and national implementation process, incorporating community and stakeholders perspectives, and presents findings from 655 couples in our initial pilot to refine operational feasibility and workflow.

## Methods

### Study design and governance

Singapore’s demographic and cultural diversity provide a unique landscape to pilot and evaluate reproductive carrier screening. The population includes Chinese (74.3%), Malay (13.5%) and Indian (9.0%) ancestries, with diverse religious beliefs including Buddhism, Islam, Christianity, and Hinduism. There are ∼34,000 births per year in Singapore(14) and since the implementation of thalassemia carrier screening in 1993, affected births with thalassemia have reduced from 7-9 annually to only rare occurrences, demonstrating its efficacy(7). Reproductive carrier screening beyond thalassemia is neither routinely offered, nor embedded in mainstream healthcare practice. Although available through commercial providers, uptake is limited due to cost barriers and low awareness, highlighting the need to expand access.

This study is a prospective, longitudinal study with multiple aims: 1. Expand reproductive carrier screening to 39,000 couples across Singapore for genetic conditions relevant to the population; 2. Assess uptake of reproductive carrier screening, provide increased risk couples with reproductive options and to review reproductive decisions; and 3. Explore multi-cultural population perspectives towards carrier screening implementation. Ethics approval was obtained by the SingHealth Central Institutional Review Board (2019/2243) and all participants in this study have provided written informed consent.

During development of this program several working groups were established, meeting fortnightly from May 2023 until present, to support the delivery of key components including gene panel design, laboratory assay development, bioinformatics optimisation and variant analysis, information resources, clinical implementation and acceptability assessment. The program is governed by a scientific advisory committee which convenes quarterly to review strategic direction, research activities and implementation progress.

### Carrier screening panel

The selection of genes comprising the carrier screening panel has been previously reported(13). Briefly, genomic data from 9,051 participants of Chinese, Indian and Malay genetic ancestry were analysed to identify pathogenic variants in 4,143 disease genes. Inheritance and associated conditions at a variant level were determined to calculate the frequencies of autosomal recessive conditions in Chinese, Indian, and Malay populations. Gene selection prioritised severe paediatric diseases and population relevance, with review from genetics specialists and key stakeholders such as obstetricians and gynaecologists, healthcare policy advisors and funding organisations. Conditions from the local genetics registry were also considered resulting in a panel of 112 genes associated with severe paediatric autosomal recessive disease. In Singapore, genetic testing in asymptomatic individuals that implies personal health risks can have insurance implications and must be clearly discussed in pretest counselling. To minimise the possibility that insurance concerns may deter participation, variants with implications for heterozygotes associated with recessive, X-linked and dominant conditions were excluded.

### Participant outreach

This carrier screening program is being implemented in two phases. The pilot phase is currently offering testing to 1,000 couples at Singapore Health Services, the largest public healthcare group. This will be followed by expansion to include the other two healthcare clusters in Singapore, reaching 39,000 couples with no out-of-pocket costs over a three year period. Information regarding this carrier screening program has been disseminated to healthcare professionals through multiple talks at conferences, forums and seminars, and to the public through media launches commencing in September 2024 (Supplementary Appendix). Recruitment pathways have been established through genetics and obstetrics clinics. In the pilot phase, recruitment is available to non-pregnant married couples where at least one partner is a Singaporean or permanent resident and has been a past or existing patient at the hospital site. Any couple with a known carrier risk, from either previous carrier screening or having an affected child, are eligible to participate as this panel will assess their risk for other genetic conditions previously not screened. However, counselling provided through this program will not address their previously identified carrier risks.

### Education platform

Participants receive an information brochure designed by our clinical genetics team with consultation by healthcare professionals at the tertiary hospital. This brochure is available in the four national languages, English, Chinese, Malay and Tamil, and contains a QR code that links to collect information to assess for eligibility (Supplementary Figure S1). Once completed and deemed eligible by our clinical team, they are directed to Genetics Adviser(15), an online education platform tailored to this screening program (https://carrierscreening.sg/). This education tool was designed over 15 months with the clinical team, comprising of geneticists and genetic counsellors, and the Genetics Adviser team to develop an education interface to ensure culturally relevant content. It includes vignettes relevant to our local context using genetic conditions and couples with ancestries representative of Singapore’s diverse population, as well as a modified decision aid(16) to guide couples in considering the implications of participating and understanding potential results. After completing the pre-test education modules on Genetics Adviser, couples are directed to a second form, capturing demographic, personal, and family history information. A brief note summarising the key information about the program is presented at the end of the form, which the couples acknowledge before submission. The clinical team reviews this information and schedules an appointment at the hospital for genetic testing consent and sample collection. A genetic counselling appointment can be also arranged for couples who would prefer to receive pre-test information in person or have any additional questions by contacting the clinical study team directly. Key counselling elements are reinforced during the genetic test consent and sample collection appointment. Couples are encouraged to attend the appointment together.

### Sample collection and analysis

In the current pilot phase, 3-5ml blood is collected for sequencing while buccal swab protocols for alternate collection are being developed for the next phase. Extracted DNA undergoes library preparation using our customised carrier screening panel and are then sequenced on a NextSeq instrument (Illumina). The sample sequencing and variant analysis occurs inhouse. The raw sequencing data are aligned to the human reference genome (GRCh38) using the BWA-MEM2(17) aligner algorithm. This is followed by read sorting and marking of duplicate reads. Processed read alignments are then analysed using the Genome Analysis Tool Kit (version 4.1.9)(18). Read alignments for each sample undergo base quality score recalibration, following by variant calling using the HaplotypeCaller algorithm. These variant calls across the entire gene are then annotated using Variant Effect Predictor (release 109.3)(19). Apart from annotating the consequence of these variants (for example, synonymous, nonsynonymous, frameshift, etc), various knowledge bases are used to annotate the variants including population allele frequency databases (gnomAD4.1)(20), local population frequencies(12), computational predictions (e.g. REVEL(21), AlphaMissense(22)) and clinical variants(23). Copy number variants for the genes in this screening panel are identified using ExomeDepth(24), and *SMN1* and *HBA1/2* copy number variants are called using additional in-house developed algorithms by the bioinformatics team. Quality control metrics for each sample are derived from Genome Analysis Tool Kit Depth of Coverage (version 4.1.9) and Qualimap (version v2.2.1)(25) analyses. Samples are required to meet minimum QC thresholds, including at least 1 million mapped reads, mean coverage of ≥50×, ≥99% of high-confidence target regions covered at ≥20×, and a duplication rate <10%. At the variant level, a minimum coverage of 20x is required, and hard filtering was applied following the thresholds recommended by GATK. Single nucleotide variants and small deletions/ insertions are curated according to ACMG/AMP (American College of Medical Genetics/Association for Molecular Pathology) guidelines(26), where a point-based variant classification category(27) is assigned according to specific criterion review information available(28).

The pipeline is designed to flag couples that have variants (both single nucleotide and copy number variants) occurring in the same gene and only these variants are then curated. The genes in this panel were selected for their association primarily with shortened lifespan or severe intellectual disability. In addition, phenotype severity and penetrance is assessed for each variant and variant combination specific to the couple. Couples are reported as increased risk only when both carry likely pathogenic and pathogenic variants that together infer severe disease. For example, *CFTR* variants associated with cystic fibrosis are reported whereas those associated with milder conditions such as congenital bilateral absence of the vas deferens are classified as low risk. Clinical impact of variants is reviewed by the bioinformatics, variant curators and clinical team. When further input is required, these variants are presented to the scientific advisory committee for discussion and consensus on the expected clinical phenotype association.

### Results provision

Reproductive risk for each couple is reported as either low or increased risk. Results for couples at low risk of having a child with a genetic condition associated with 112 genes screened are returned by teleconsult. They receive their low-risk result report and summary of test limitations by encrypted email and are redirected to Genetics Adviser for post-test genetic counselling which summarises the implications of their results. Results for couples found to be at increased risk of having an affected pregnancy are returned through an in-person genetic counselling appointment to discuss the results of their test report and available reproductive options. These include pre-implantation genetic testing (with reimbursement for one cycle), prenatal diagnosis via chorionic villus sampling or amniocentesis, gamete donation, or adoption. Another blood sample is collected to clinically validate findings (Figure 1).

**Figure 1.**
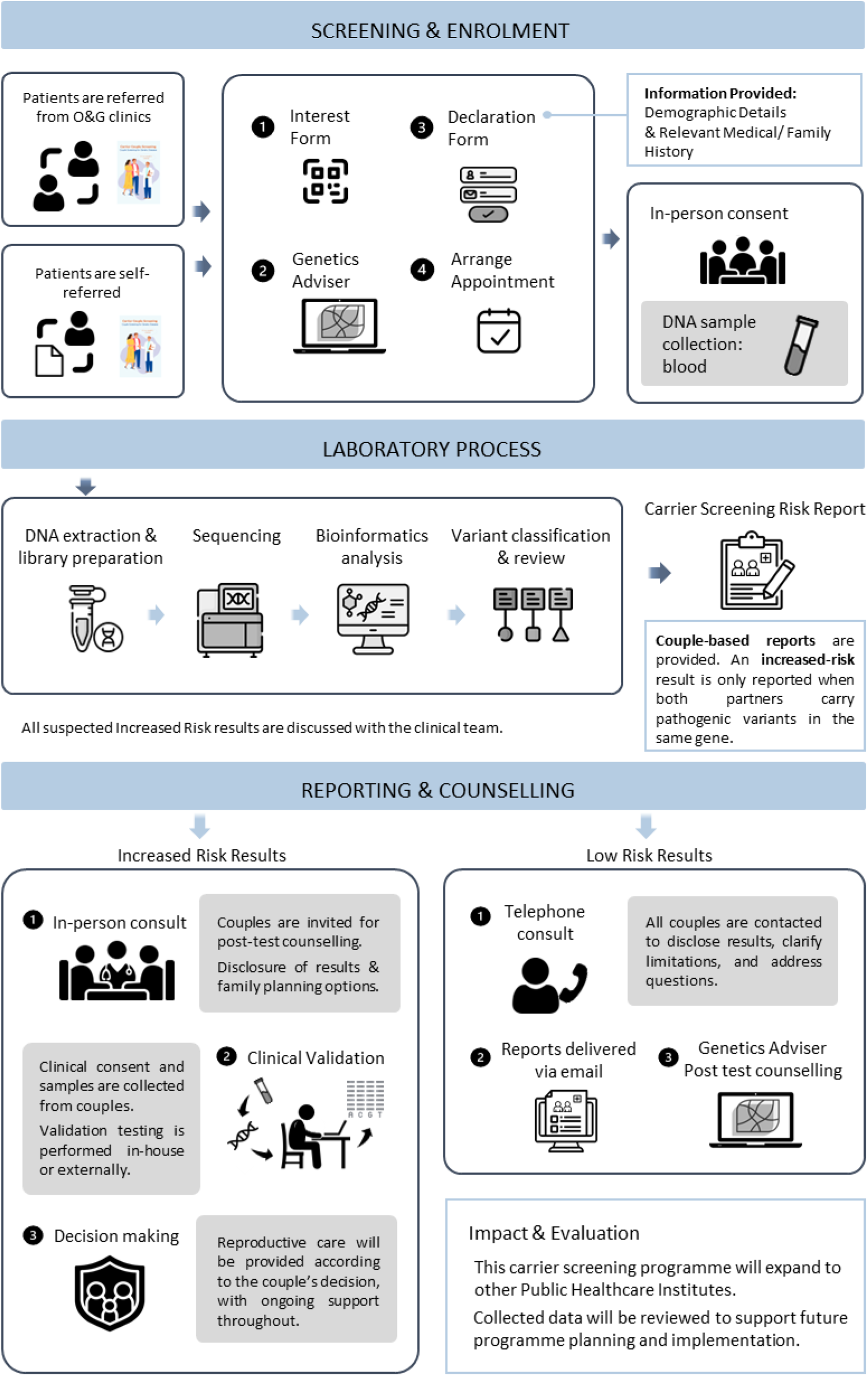
Overview regarding participant journey from awareness to results provision.

### Stakeholder perspectives in carrier screening implementation

Given the diverse ethnic and religious background of the population, we engaged multiple stakeholders, including couples of reproductive age (community members), healthcare professionals and religious leaders.

#### Community Questionnaire

A cross-sectional questionnaire was developed based on previous literature(29–32), then piloted with community members to assess the clarity and cultural appropriateness. Individuals of reproductive age were recruited through Singapore Health Services, the general community and participants who consented to our carrier screening program. Completion of the questionnaire was voluntary and independent of enrolment to program involvement. The questionnaire explored multiple domains regarding interest and attitudes toward carrier screening participation. Demographics, reproductive and awareness variables were collected to explore associations between respondent characteristics and their awareness, attitudes, and intended reproductive behaviours related to carrier screening (Supplementary Appendix).

#### Healthcare Professional Questionnaire

In parallel, healthcare professionals working in genetics, obstetrics, and related specialties at Singapore Health Services completed a questionnaire assessing their perspectives on carrier screening implementation in clinical practice. The questionnaire was developed from literature describing the integration of carrier screening into healthcare systems(33–35), and explores clinician knowledge, attitudes, implementation feasibility and screening condition preferences with professional role and genetic testing experience collected as potential influencing factors (Supplementary Appendix).

#### Religious leader engagement

We also engaged with religious leaders representing Islam, Buddhism, Christianity, Hinduism, Catholicism, Sikhism and Judaism communities to describe the carrier screening program aims and conditions to be screened. We shared our implementation pathway and discussed reproductive options available for increased risk couples planning a family such as preimplantation genetic testing, antenatal testing, termination, gamete donation and adoption.

### Statistical analysis

All statistical analyses were performed with the SciPy1.0(36) and statsmodels(37) packages. Descriptive analyses were used to describe the sociodemographic characteristics of the respondents, their interest and intentions for participating in carrier screening and views towards reproductive options. The relationship between demographic categories, awareness and interest level was investigated using the chi-square test. Ordinal logistic regression was used to compare scaled responses.

## Results

### Carrier screening implementation

#### Recruitment

As shown in Figure 2, 1,619 couples have registered their interest to date. In this pilot phase, 467 couples not meeting eligibility criteria (married, Singaporean or permanent resident, current or former patients of the hospital site and not pregnant) have been placed on a waiting list for the program’s next phase. Of the 1,116 deemed eligible to participate, 799 (69%) couples completed the education module, a further 55 were deemed ineligible, and appointments were arranged for 744 couples to provide consent and samples. There were 18 (2.4%) couples that declined testing either prior or during the consent appointment and 70 have appointments pending. No couples requested an additional in person pre-test genetic counselling appointment. Since sample collection, only one couple has requested to withdraw and 10 couples have reported being pregnant, prompting expedited processing of their results. Of the 1,097 eligible couples, with 522 excluded due to ineligibility, 60% (n=656) have provided consent for carrier screening to date.

**Figure 2.**
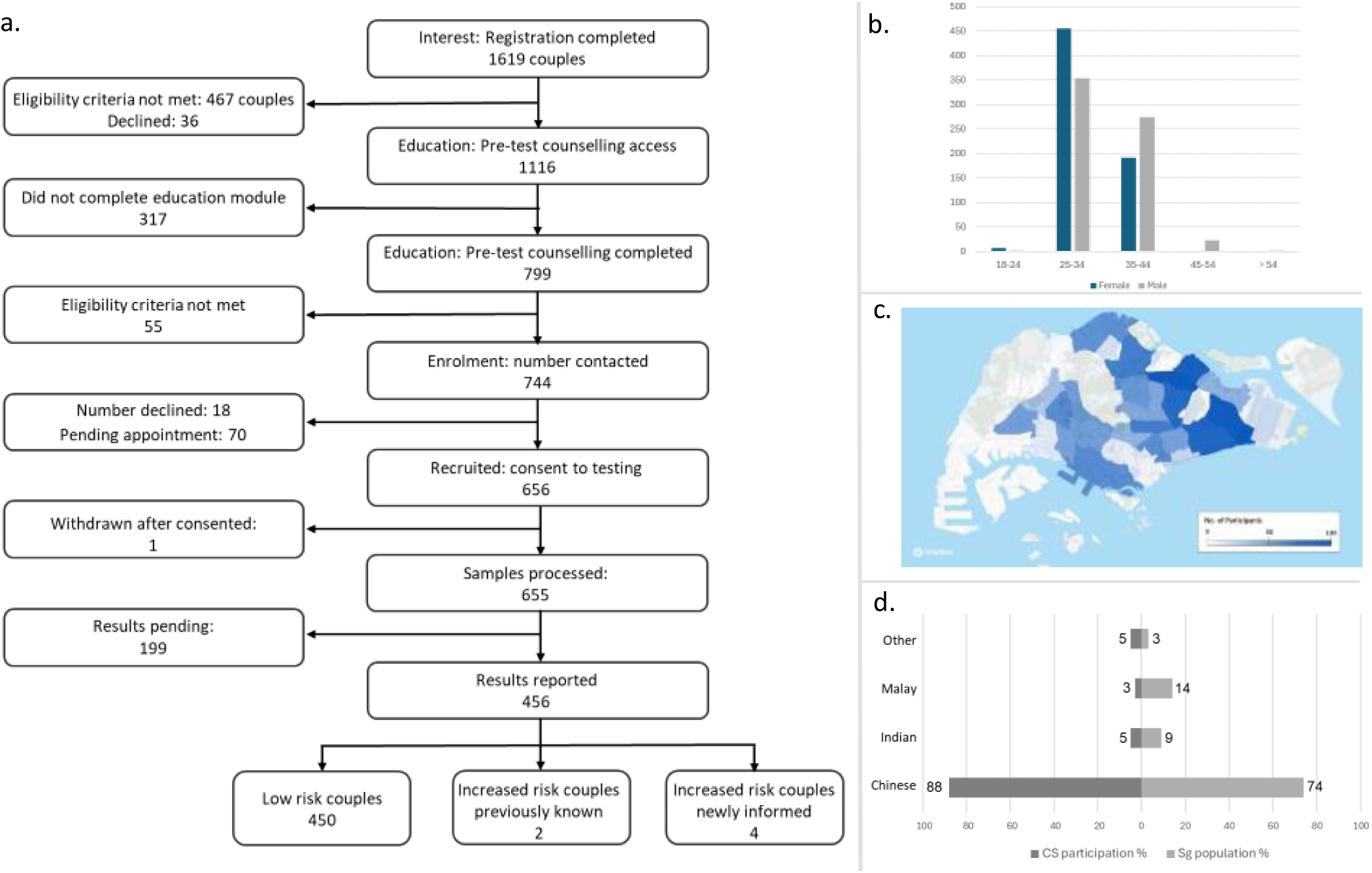
Participation of carrier screening (CS) overview. a. Participant engagement in the carrier screening program b. Age of participants who consented to participate and have samples processed (n=1310, 655 couples) c. Residential distribution of consented participants across Singapore (n= 1310). d. Percentage of consented participants by self-reported ancestry (CS participation) in comparison to the Singaporean (Sg) population (n=1310)

#### Results delivery

Of the 656 couples that proceeded with carrier screening, 1 couple withdrew. Consented participants reside across all regions in Singapore (n=655), with participant age ranging between 21 to 59 years (median: 33.5 years) and ancestry self-reported as Chinese (88%), Indian (5%), Malay (3%) and Other (5%) (Figure 2, Table 1). There are 456 (69.4%) couples who have received their results with 450 (98.6%) couples found to be at low risk of having a child affected by a genetic condition included the 112 gene panel. The remaining six (1.4%) couples were assessed at increased risk, two of these couples were aware of their carrier risk prior to enrolment (HbH disease, OMIM 613978 and glycogen storage disease type II, OMIM 232300) while the remaining four had no previously known risk according to family history or consanguinity. Among these four couples, two couples were found to be carriers *HBA1/2* associated with alpha thalassemia (OMIM 604131) and HbH disease (OMIM 613978). The remaining couples were found to be carriers of *WRN* (Werner syndrome, OMIM 277700) and *CEP290* (*CEP290*-related ciliopathy, OMIM 610188) (Table S1). One couple became pregnant, and subsequently proceeded with prenatal testing for *CEP290*-identified variants which showed that the fetus was unaffected.

**Table 1.** Demographics of participants who have consented to carrier screening.

| **Characteristics** | **Total**<br>**n= 1310** |
| --- | --- |
| *Gender* |  |
| Female | 655 (50%) |
| Male | 655 (50%) |
| *Ancestry* |  |
| Chinese | 1148 (88%) |
| Indian | 68 (5%) |
| Malay | 33 (3%) |
| Other | 61 (5%) |
| *Age* |  |
| 18-24 years | 9(7%) |
| 25-34 years | 809 (62%) |
| 35-44 years | 466 (36%) |
| 45-54 years | 24 (2%) |
| Over 54 years | 2 <1%) |
| *Results* |  |
| Increased Risk | 12 (0.9%) |
| Low Risk | 898 (69%) |
| Pending | 398 (31%) |

### View towards carrier screening implementation

#### Community perspectives

##### Community demographics

In total, 1002 community members completed the questionnaire. All respondents were of reproductive age, 557 (58%) were female, 821 (82%) were married, 292 (29%) have children and 636 (63%) indicated wanting children or more children. The majority of the responses derived from individuals of Chinese ancestry (76%), followed by Malay (12%), Indian (7%) and Other (4%) ancestry. Regarding religious affiliation, Buddhist (26%), Christian (24%) and Muslim (16%) affiliations were the most common, whereas 25% did not affiliate with a religion. There were 266 (26%) respondents that had consented to this carrier screening program which was voluntarily offered at time and the remaining 736 (75%) were approached from the general community (Table S2).

The responses from the community members were mapped across the program journey, starting with awareness as the first stage of program interaction. We then explored interest, participation intentions and influencing factors, alongside barriers that may prevent participation. Finally, we assessed the perceived implications of participation for the individual and their views towards reproductive choices (Figure 3).

**Figure 3.**
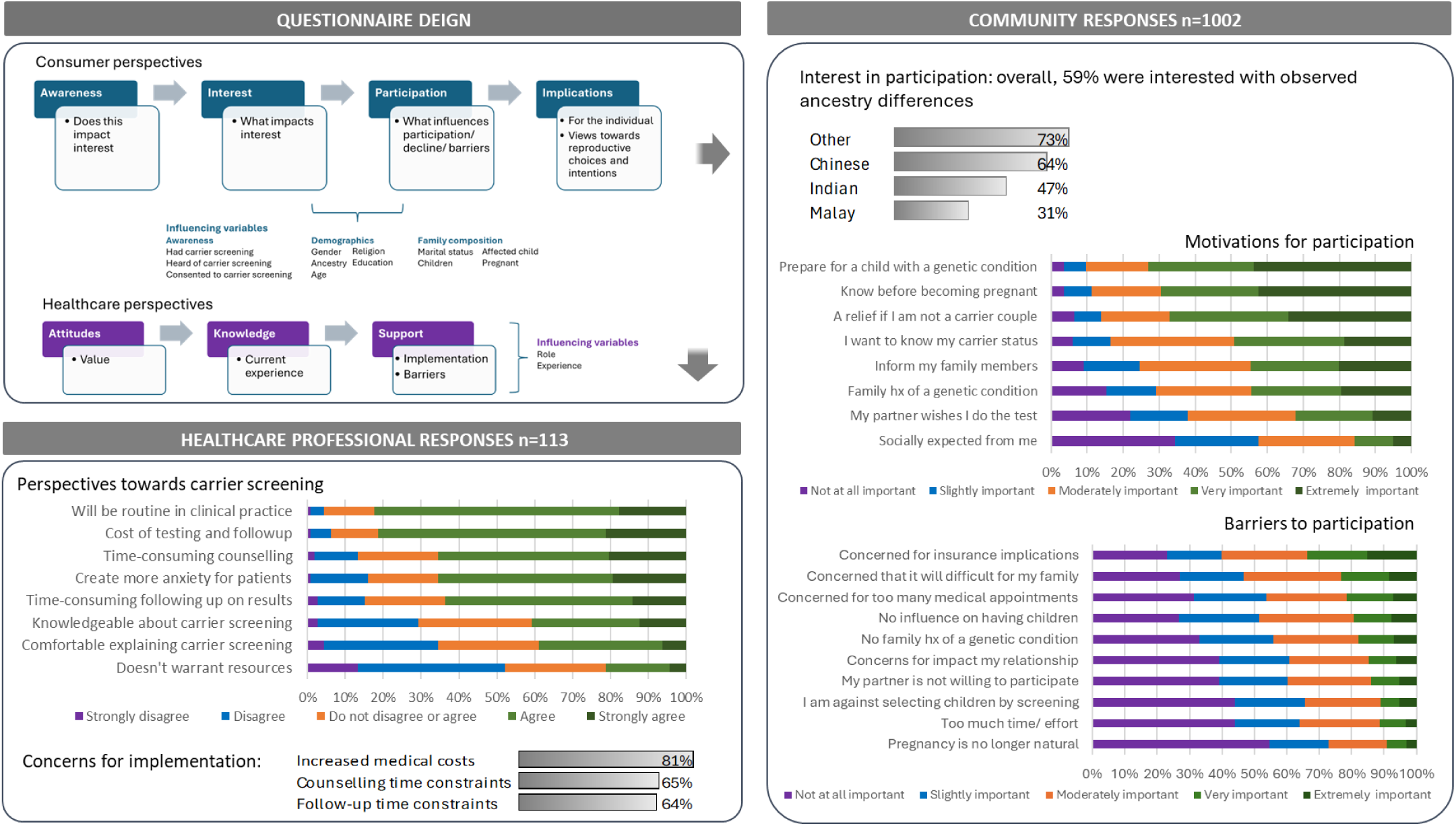
Questionnaire analysis and responses overview.

##### Awareness, interest and participation

Overall, 43% of respondents had heard of reproductive carrier screening, though only 9% had previously undergone testing. As shown in Table S2, awareness differed across demographic groups, with higher awareness among females (*p*= 0.0228), those with a higher education (*p*= 0.0001), respondents of Chinese ancestry (*p*= 0.0001), and individuals with either no religious affiliation or identified as either Taoist, Christian, or Buddhist religions (*p*= 0.002); those not pregnant (*p*= 0.0129) Those who consented to carrier screening also had a higher prior awareness (*p*= 0.0016). Respondents expressed high interest in participating in carrier screening, with 59% reporting they were extremely or very interested (Figure 3), however there was notable variation observed across ancestry (*p*= <0.0001), religion (*p*= <0.0001), pregnancy status (*p*= <0.0001), and reproductive intentions (*p*= <0.0001) (Table S3). Respondents of Chinese ancestry, Buddhist and Christian affiliation and those planning to have children showed the highest interest, while pregnant individuals were less interested. Those that had consented to participate showed more interest than respondents that had not (Figure S2a). Prior awareness of carrier screening was associated with a greater intent to participate with 69% extremely or very interested (*p*= <0.0001). Those who had prior awareness of carrier screening and had consented to this program showed the greatest interest, with 84% extremely or very interested, followed by no prior awareness yet had consented the program (77%, *p*= 0.176) (Table S3).

Motivations to participate considered extremely or very important were the ability to prepare for a child with a genetic condition (73%), knowing risk before pregnancy (69%), and reassurance if not a carrier (67%) (Figure 3). Motivation for preparation tended to be higher among younger individuals (*p*= 0.00045), those with higher education (*p*= 0.0002), and those considering children (*p*= <0.0001). Social influences were regarded as less important yet varied by religion and pregnancy status (Table S4). Concerns for insurance implications was viewed as most important consideration to decline carrier screening (33%) (Figure 3), shared by more Chinese respondents in comparison to Malay (*p*= 0.01900) (Table S4). Other reasons to decline carrier screening included concerns that it would be difficult for the family (23%) or having too many medical appointments (22%) (Figure 3).

##### Anticipated implications

Upon receiving an increased risk result, overall respondents strongly agreed or agreed they would feel relieved that further testing could be performed (86%) and informed (75%) (Figure S2b) and these sentiments were consistent across all demographics (Table S4). However, 67% also strongly agreed or agreed they would feel anxious (Figure S2b), which was slightly more observed among pregnant respondents (78%,) compared to those not pregnant (72%, *p*= 0.349), Indian (74%) in comparison to Chinese (67%, *p*= 0.185) and Malay (58%, *p*= 0.0049) respondents and females (73%) compared to males 58%, *p*= <0.0001) (Table S4).

Regarding being at increased risk and pregnant, most strongly agreed or agreed that these results would be important for preparation (87%) and would consider further fetal assessment (85%) (Figure S2c). There were 72% of respondents that strongly agreed or agreed with considering termination, this response was shared more by Buddhist (79%, *p*= <0.0001) and Christian (73%, *p*= 0.00208) or those with no affiliation (73%, *p*= <0.0001) in comparison Muslim (58%) respondents. Around half of all respondents (51%) agreed or strongly agreed that they would consider not having any more children if found to be at increased risk and pregnant which was shared by 69% of respondents not wanting children in comparison to those wanting children (43%, *p*= <0.0001) (Figure 2c, Table S4).

#### Healthcare professional perspectives

##### Healthcare professional demographics

Among the 113 healthcare professional responses that completed the questionnaire, responses comprised of 52 (46%) doctors, 44 (39%) nurses, 13 (12%) allied healthcare professionals and 4 (4%) genetics counsellors. Over half (56%) indicated they offer genetic testing in their clinical practice and of these, 29 (46%) were involved in offering reproductive carrier screening. Only 16 (14%) respondents worked in a dedicated genetics service (Table S5).

##### Experience with carrier screening

Overall healthcare professionals indicated mixed but generally positive view to carrier screening. Most respondents (83%) strongly agreed or agreed that eventually carrier screening will become routine in clinical practice. However, only 20% thought that the associated risks to couples were substantial enough to warrant dedicated resources. Many highlighted concerns about increased medical costs for patients (81%) as well as time commitments regarding pretest counselling (65%) and results follow-up (64%). Patient anxiety was also a notable concern (65%) (Figure 3). Only 40% strongly agreed or agreed with feeling knowledgeable and 39% felt comfortable in explaining carrier screening to patients. Even among those currently offering carrier screening, only 39% reported feeling knowledgeable (Table S6).

##### Perspectives on offering screening

The majority strongly agreed or agreed that pre-test counselling is required for all couples considering carrier screening (87%) and that it requires informed consent (96%) that could be provided by appropriately trained healthcare professionals (83%) in addition to genetics specialists (48%) or by video with optional pretest counselling (70%) (Figure S3a). There was strong agreement or agreement that carrier screening should be included in preconception counselling (87%). For high risk pregnancies, for example relevant family history (90%), consanguinity (87%), or an abnormal first trimester pregnancy screen (73%), there was strong support for carrier screening to be offered. However only 58% agreed that carrier screening should be offered to all couples considering a pregnancy (Figure S3b).

##### Personal choices of healthcare providers regarding carrier screening

When asked if they would personally participate in carrier screening, 31% of healthcare professionals strongly agreed or agreed they would decline preconception carrier screening due to concerns regarding increased anxiety, rising to 43% if offered during pregnancy. There was a preference to screen larger over a smaller panels (87%) if there was no increase in cost. If found to be at increased risk, the majority indicated they would opt for further testing such as prenatal diagnosis (90%) or pre-implantation genetic testing (88%) (Figure S3c).

#### Religious leader engagement

We consulted religious leaders representing Islam, Buddhism, Christianity, Hinduism, Catholicism, Sikhism and Judaism to outline the program and review reproductive options available for increased risk couples. All religious leaders were supportive of carrier screening being offered and with varied views towards the reproductive options and acceptance within their community. Engagement extended with Islamic leadership leading to further research by Islamic scholars reviewing Islamic text and presenting their findings at Fatwa Lab symposium in June 2025. This was followed by issuance of a Fatwa supporting carrier screening for Malay Muslim couples(38) (Figure S4).

## Discussion

Analysis of local genomic data showed significant gaps in ancestry representation in publicly available databases and commercial panels, presenting an opportunity to develop a screening program inclusive of Asian populations. Here we describe the development of a pilot nationwide carrier screening program for an Asian cohort, addressing the paucity of such initiatives beyond European ancestries. We report the experience of screening 456 reproductive couples using a customised 112 gene panel associated with over 80 severe paediatric recessive conditions prevalent in the target population. These early implementation insights and questionnaire responses are guiding development towards an expanded population-based carrier screening program in Singapore.

Since initiation, 1,619 couples have registered interest, with broad residential distribution, and the majority of those eligible continued with pretest education. To date, screening uptake, defined by couples who registered interest, deemed eligible and then consented to carrier screening, is 60% however this may decrease as the program continues to scale. In comparison, a recently reported nation-wide implementation program reported uptake of 49.9%(9) which is more closely aligned to other studies(39–41), however parameters defining uptake may vary. So far, few couples (5%) have declined participation or requested additional genetic counselling after receiving program information, suggesting that the online pretest education platform and the consent appointment can support decision making regarding participation. Although positive outcomes have been reported for digital tools used in carrier screening pretest counselling(42), further research is required to analyse their impact in this setting. Among the 456 couples screened, the detection of newly increased reproductive risk in 0.9%. While it is possible that our pilot may be self-selecting for individuals, especially for a thalassemia history, we identified two couples with less common diseases, highlighting the potential of expanded carrier screening beyond thalassemia. The identification of previously unknown risks for Werner syndrome and *CEP290*-related ciliopathy suggests the utility of expanding carrier screening beyond the traditional approach limited to thalassemia in Singapore.

Complementing these implementation findings, overall interest from the community questionnaire data was also high (59%), which was either similar(31, 43, 44) or higher(29, 32, 45) than other comparable studies. However, there were marked demographic differences regarding awareness and willingness to participate. Respondents were representative of the major ancestry and religious groups in Singapore, the majority are of reproductive age, married or intending to have children, and therefore have provided key insights towards implementation. Despite, thalassemia carrier screening being routinely offered during pregnancy, less than half of respondents (43%) had heard of carrier screening. Differences were observed across many demographic groups highlighting how sociocultural and educational factors could shape awareness of genetics services. Notably, individuals with higher education, those not pregnant and respondents of Chinese ancestry demonstrated higher awareness.

These patterns also aligned with interest in participation and indicate that greater awareness is associated with a higher intent to participate. Respondents of Chinese ancestry indicated strong interest, consistent with their high engagement in the carrier screening program. In contrast, respondents of Indian and Malay ancestry reported lower interest, despite the availability of carrier screening information brochures available in the four national languages and disseminated through the tertiary hospital. Similarly, religion appears to play a role in interest despite all religious leaders expressing support. Respondents with religious beliefs have been shown in several studies to be less interested(29, 32, 43), yet in one Belgian study there was no such association(46), however diverse religious groups were not defined. In our setting, engagement with the Islamic community extended to comprehensive research regarding implications for reproductive decisive making and has resulted in a recently issued Fatwa encouraging participation. However, interest among Muslim respondents was lower and therefore reviewing engagement from this community following its release will be valuable.

Interest was higher amongst those not pregnant, which contrasts with existing practice where thalassemia is offered during pregnancy. Motivations around program participation placed importance on preparation at preconception and this was emphasised by those not pregnant or planning a pregnancy. Concerns about insurance implications emerged as a prominent barrier, particularly among Chinese respondents and those with higher educational attainment and has also been cited in a previous study (47). Genes currently being screened are associated with autosomal recessive conditions and therefore have minimal implications for carriers and insurance applications, highlighting an area for further education. Collectively, these findings align with current recommendations(8) and previous studies (29, 30, 32, 43) that culturally appropriate education at preconception should continue for program engagement and equitable uptake.

Although the majority of healthcare professionals agreed that carrier screening will become routine in clinical care, significant gaps in preparation for implementation were apparent. Concerns regarding increased medical costs and counselling time were predominant with only half agreeing that the associated reproductive risks were sufficient to justify prioritising resources. Similar challenges regarding time and resource burden have been documented in previous studies involving healthcare professional perspectives(33, 34, 48). Notably, despite more than half of respondents offering genetic testing in their current practice, less than half reported confidence and knowledge specifically in carrier screening, even amongst those offering it, highlighting the need for additional education and support. Only 58% of respondents indicated that carrier screening should be offered to all reproductive couples, despite professional guidelines recommending this approach(8). This is consistent with a healthcare practice review reporting views that offering testing should be patient led(35) rather than routinely offered. Education tools, such as the adoption of Genetics Adviser to complement pre and post-test counselling, are recognised to effectively improve patient understanding while reducing counselling time for providers(34, 49). In addition to supporting healthcare provider education regarding carrier screening, increasing awareness of our implementation and education processes may alleviate concerns about information provision and support expansion.

Although prospective parents may express high interest in participation, uptake is typically lower, known as the Intention-Behaviour gap, where the influence of both internal and external factors can prevent self-reported intentions correlating with observed behaviour(50). In this study, both interest and uptake were comparable, possibly due to no out of pocket costs. However, couples are encouraged to register and attend an in-person appointment together which may pose practical barriers. Exploring which aspects of the program encourage uptake will be valuable for refining future implementation. Further work will also involves a health technology assessment to evaluate resource utilisation and cost per increased-risk couple identified according to reproductive preferences. With declining sequencing costs, whole genome sequencing may enable a “Sequence Once, Analyse Repeatedly” approach, allowing analysis for recessive disease risk while supporting broader precision medicine applications such as pharmacogenomics.

## Conclusions

Our experience, together with insights from previous studies, indicates that sociodemographic factors and implementation models vary substantially across communities and may not directly translate into local contexts. This highlights the importance of reviewing these influences within the target population to identify context-specific determinants to awareness, interest and engagement. Collectively, our early findings support the feasibility of integrating population-level carrier screening into routine reproductive care as precision medicine efforts expand in Singapore, provided that practical challenges are addressed through equitable outreach and professional training. Ongoing follow-up will inform optimisation of counselling needs and support pathways for reproductive couples.

## Abbreviations

ACMG: American College of Medical Genetics and Genomics
ACMG/AMP: American College of Medical Genetics/Association for Molecular Pathology

## Declarations

## Ethical approval and consent to participate

Ethics approval and consent from participants consenting to take part in the carrier screening program and questionnaires was obtain from SingHealth Centralised Institutional Review Board (2019/2243). All experiments were performed in accordance with relevant guidelines and regulations. This research conforms to the principles of the Helsinki Declaration.

## Data availability

The datasets analysed during the current study are available in the supplementary files and further information is available from the corresponding author on reasonable request.

## Competing Interests

SSJ is co-founder of Global Gene Corp Pte Ltd and Rhea Health Pte Ltd. SSJ has received travel allowances from Illumina, Pacific BioSciences and Oxford Nanopore. DA is Medical Director at Eugene Labs. WK is co-founder of Rhea Health Pte Ltd. GB and JC are on the scientific advisory board of Rhea Health Pte Ltd. The rest of the authors declare that they have no competing interests.

## Funding

This study is funded by the carrier screening grant MCHRI/FY2023/EX/152-A207, AM/ACP-Designated Philanthropic Fund Award MCHRI/FY2023/EX/152-A207 through Temasek Foundation and AM Strategic Fund Award PRISM/FY2022/AMS(SL)/75-A137, SUNRISE SingHealth Duke-NUS AM Strategic Fund Award PRISM/FY2022/AMS(SL)/75-A137, National Precision Medicine (NPM) Phase II is supported by the National Research Foundation, Singapore (NRF) under the RIE2020 White Space (MOH-000588 and MOH-001264) funding initiative. NPM Phase III is supported by the Singapore Ministry of Health through the NMRC Office, MOH Holdings Pte Ltd under the NMRC RIE2025 NPM Phase III Funding Initiative (MOH-001734). The community questionnaire study is supported by the Social Science Research Council (Singapore) and administered by the Ministry of Education, Singapore, under its Social Sciences Research Thematic Grant (SSRC2023-SSRTG-006). This research was also supported by the Commonwealth through an Australian Government Research Training Program Scholarship [DOI: https://doi.org/10.82133/C42F-K220]. SSJ is supported by National Medical Research Council Clinician Scientist Award (NMRC/CSAINVJun21-0003) and (NMRC/CSAINV24jul-0001). WKL is supported by National Precision Medicine Programme (NPM) PHASE II FUNDING (MOH-000588).

## Author contributions

SSJ and WK were involved in the conception of the study and funding acquisition. SSJ, WK, YB, JG, GG and SC were responsible for study design. MY, SC, CS, JG and NC contributed to the day-to-day conduct of the program implementation. WK, JXT, CY, JL, WWL, HYL, SSJ, YB, SHC performed laboratory analyses, including processing samples and genomic data, and undertaking genomic data analysis and reporting. CC, JG, MY, SC, CS, JG, NC, YB and SSJ were involved with information development and dissemination. IN, PT, YHC, JKYC, GB and NG are involved with program governance. CS, MY, SC, HJT, RB, MM, JH, DA, JL, SC, YB and SSJ contributed to questionnaires design, distribution and analysis. MY, NC, SSJ, YB performed result disclosure and provided genetic counselling. YB and SSJ wrote the initial draft of the paper. All authors contributed to the critical review of the paper and approved the final version for publication.

### Acknowledgements

We are very grateful to the volunteers who have consented to take part in this program as well as community members, healthcare and religious leaders who shared their perspectives towards carrier screening implementation. We would like to thank Tai Wai Yeo, KK Women’s and Children’s Hospital and Ruifen Weng, DxD Hub for their significant contribution towards the implementation of the gene panel, Victor Effendie, National Heart Centre Singapore for the development of the reporting system, Shiqi Lim, National Heart Centre Singapore for sample and laboratory processes and Temasek Foundation for their ongoing support. We are also very grateful to Marc Clausen, Genetics Adviser, who shared invaluable experience in the development of the education platform. We would also like to acknowledge the clinical geneticists, genetic counsellors and genetics nurses at KK Women’s and Children’s Hospital for their valuable contribution in reviewing our information content.

## Supplementary Appendix

## 1. Healthcare professional education and community awareness

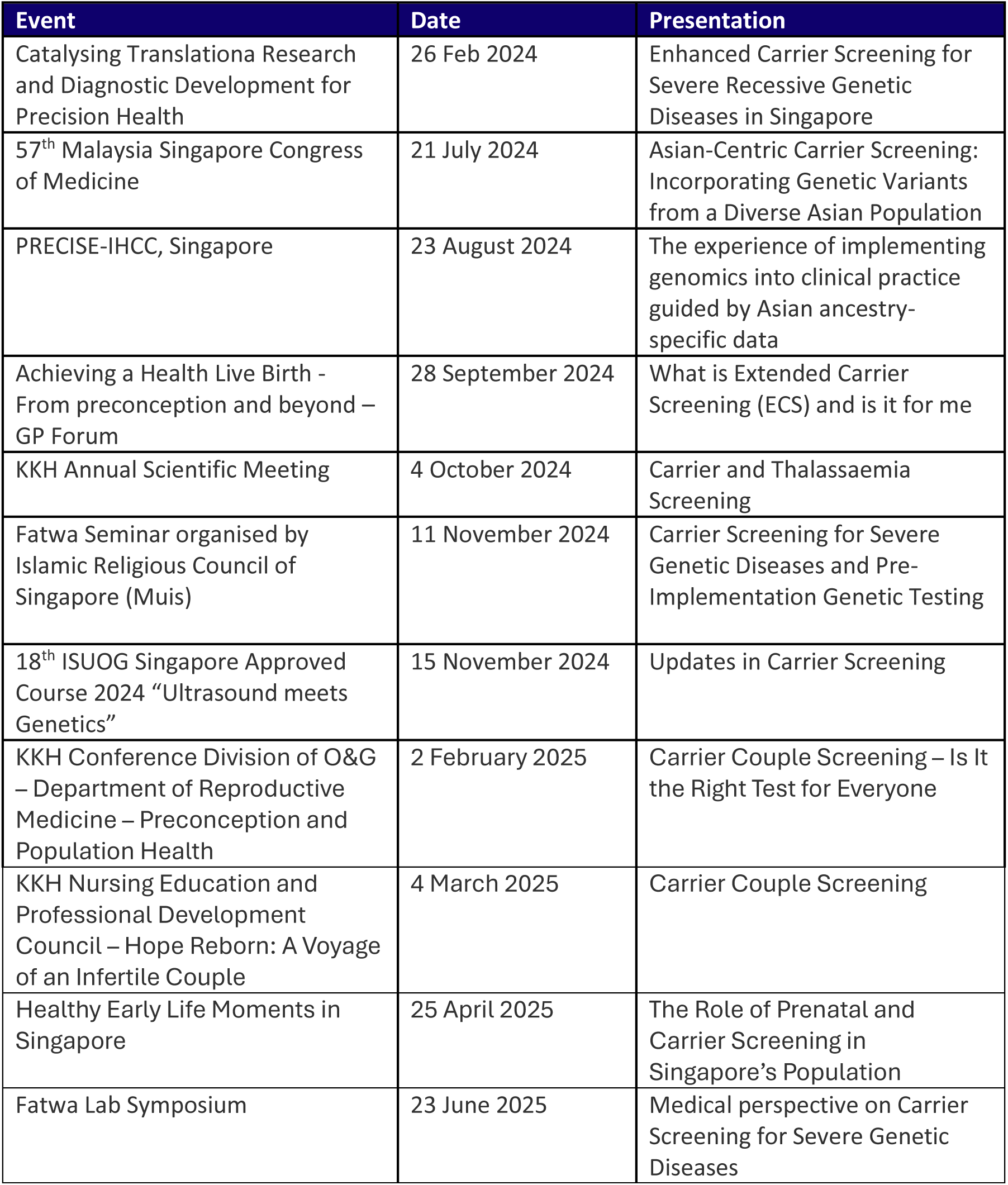

### Media launches

- Channel 8, 6.30pm (17 September 2024)
- Channel 8, 10pm (17 September 2024)
- Channel U (17 September 2024)
- Capital 95.8FM, 6pm (17 September 2024)
- Capital 95.8FM, 9pm (17 September 2024)
- Capital 95.8FM, 11pm (17 September 2024)
- Warna 94.2FM (17 September 2024)
- Oli 96.8FM (17 September 2024)
- CNA Online (17 September 2024)
- The Straits Times Online (18 September 2024)
- The Singapore Women’s Weekly (17 September 2024)
- Lianhe Zaobao Online (17 September 2024)
- Lianhe Zaobao Online (17 September 2024)
- Lianhe Zaobao (18 September 2024)
- Shin Min Daily News (17 September 2024)
- 8 World (17 September 2024)
- 8 World (17 September 2024)
- BERITA Mediacorp (17 September 2024)
- Tamil Murasu Online (17 September 2024)

The hospital was also featured in the media as follows:

- Lianhe Zaobao (18 September 2024) - Complex injuries may require specialist treatment
- Lianhe Zaobao Online (18 September 2024) - Complex injuries may require specialist treatment
- The Straits Times Online (18 September 2024) - Pioneering research to bring hope to patients with miscarriage risks and autoimmune diseases

## 2. Questionnaire development

### Participant Questionnaire

A cross-sectional questionnaire was developed to assess the acceptability of carrier screening and factors influencing participation among individuals of reproductive age, which was designed from previous literature (ref) with input from clinical genetics specialists. The questionnaire was piloted with consumers to assess the clarity and comprehensibility of the questions. Based on feedback, wording and structure were refined to ensure the language was clear, culturally appropriate, and easily understood by the target audience. Individuals of reproductive age were recruited through three channels: (1) patients attending outpatient appointments at a tertiary hospital in Singapore, (2) members of the general community, and (3) participants consenting to our carrier screening program. Eligible participants were invited to complete a questionnaire exploring attitudes and preferences toward carrier screening participation.

The questionnaire aimed to explore attitudes towards carrier screening and covered the following domains:

- Awareness and understanding of carrier screening
- Interest and willingness to participate
- Factors influencing uptake or reasons for declining
- Decision-making influences and reproductive considerations
- Perceptions of disease severity and preferences for conditions to be screened
- Implementation preferences, including cost, accessibility, and service delivery
- Preferences for receiving information and results
- Willingness to share identified or de-identified data for research

Variables collected that may influence responses include:

- Demographics: age, gender, ancestry, religion, education level, marital status, number of children, and family history of genetic conditions
- Reproductive history and intentions: current pregnancy status, plans for future pregnancy, and current children
- Awareness: whether participants had previously heard of carrier screening, had carrier screening or consented to this carrier screening program

These data allowed exploration of associations between demographic or experiential factors and participants awareness, attitudes, and intended reproductive behaviours related to carrier screening.

### Healthcare Professional Questionnaire

In parallel, a questionnaire was administered to healthcare professionals working in genetics, obstetrics, and related specialties in public healthcare institutions. The aim was to understand clinician perspectives on carrier screening implementation in current clinical practice.

The healthcare professional questionnaire was developed based on literature describing integration of carrier screening into healthcare systems and included the following components:

- Knowledge and attitudes towards carrier screening
- Awareness and prior experience with carrier screening
- Perspectives on pre-test counselling delivery and communication of results
- Feasibility and challenges of implementation in clinical practice
- Views on conditions appropriate for screening
- Personal perspectives and interest in carrier screening as consumers
- Perceived societal and research implications

Variables collected that may impact responses include professional role, years of experience, and clinical area of practice.

---

**Figure S1.**
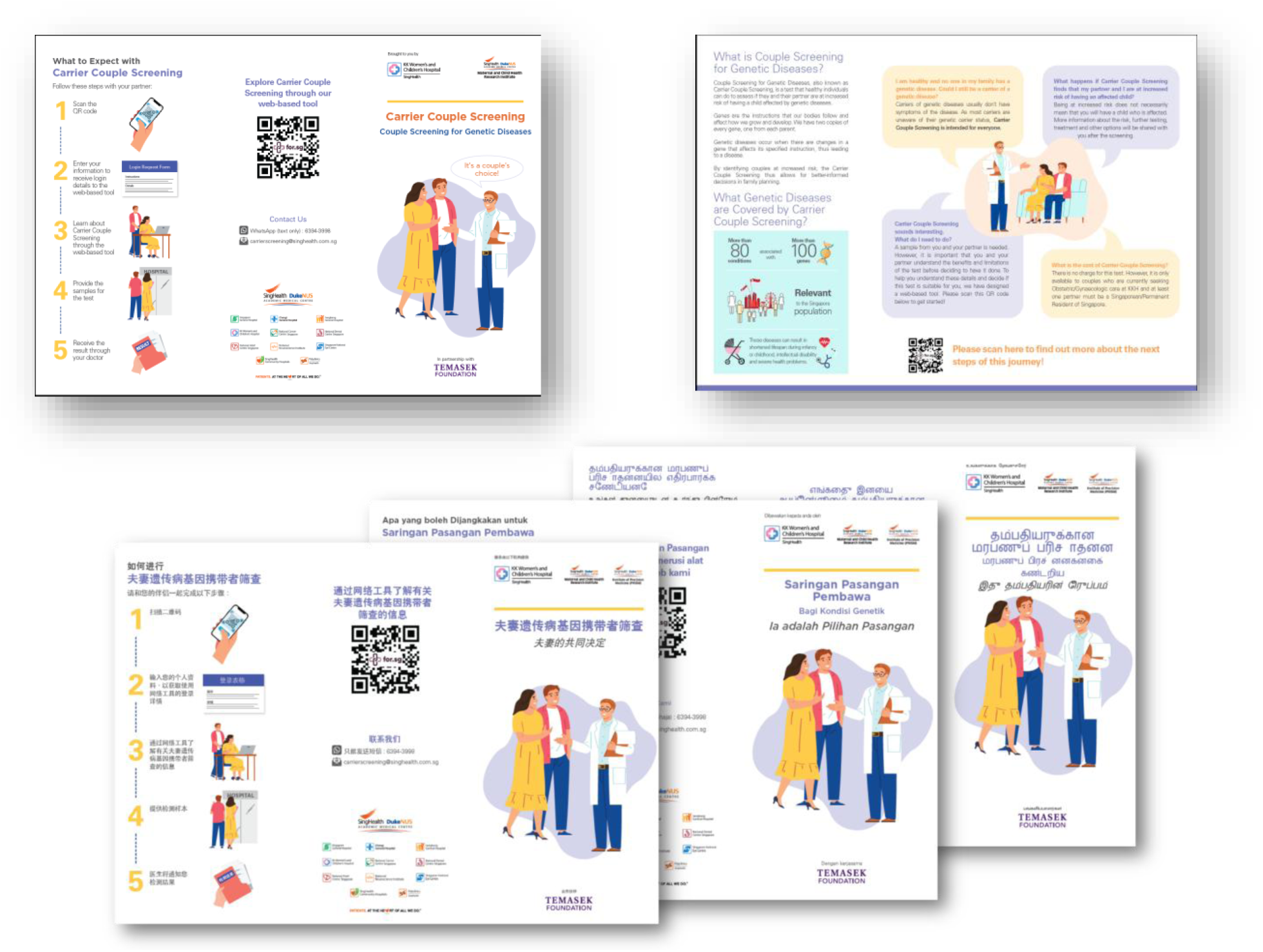
Participant carrier screening brochure.

**Figure S2.**
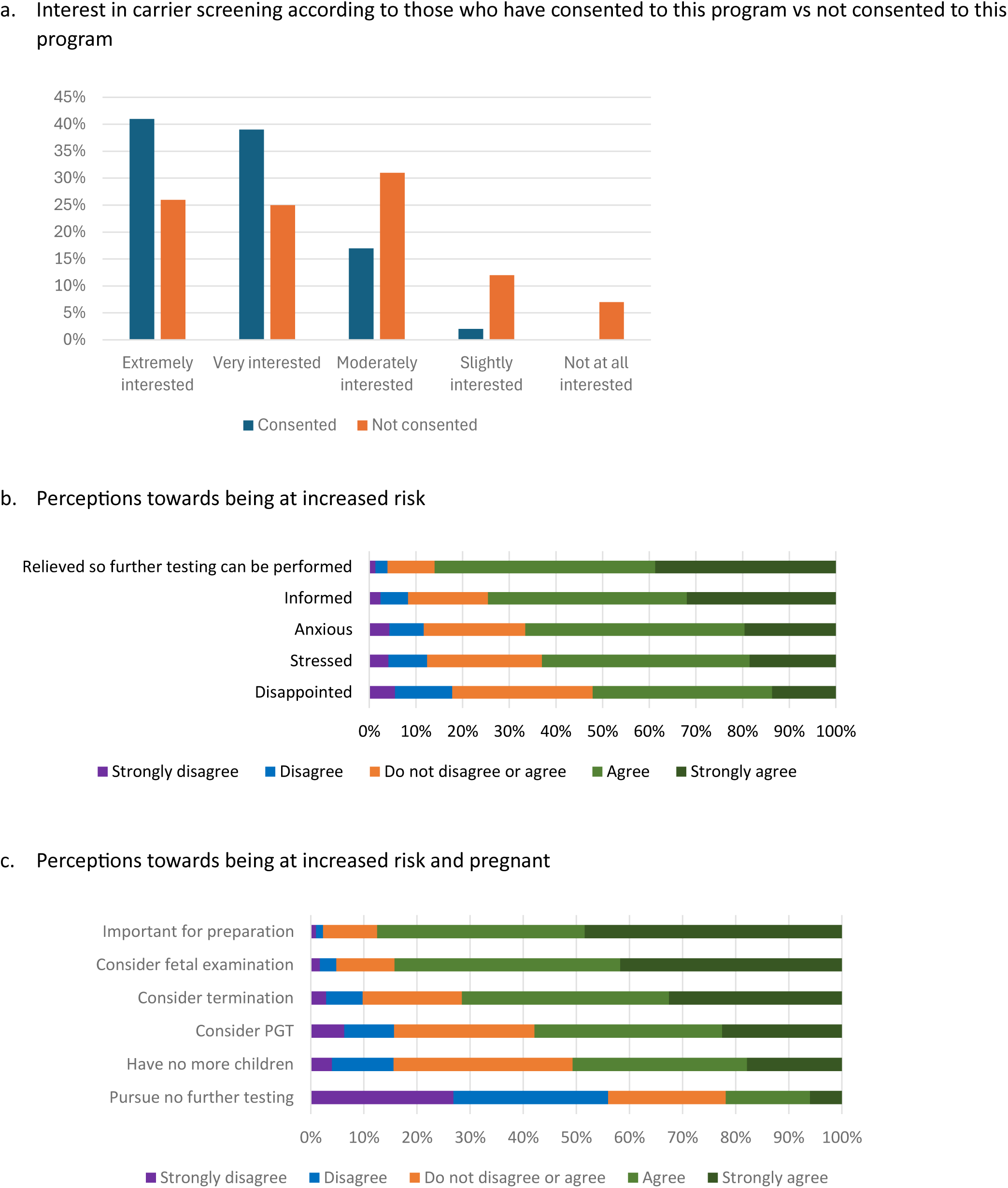
Consumer preferences towards carrier screening.

**Figure S3.**
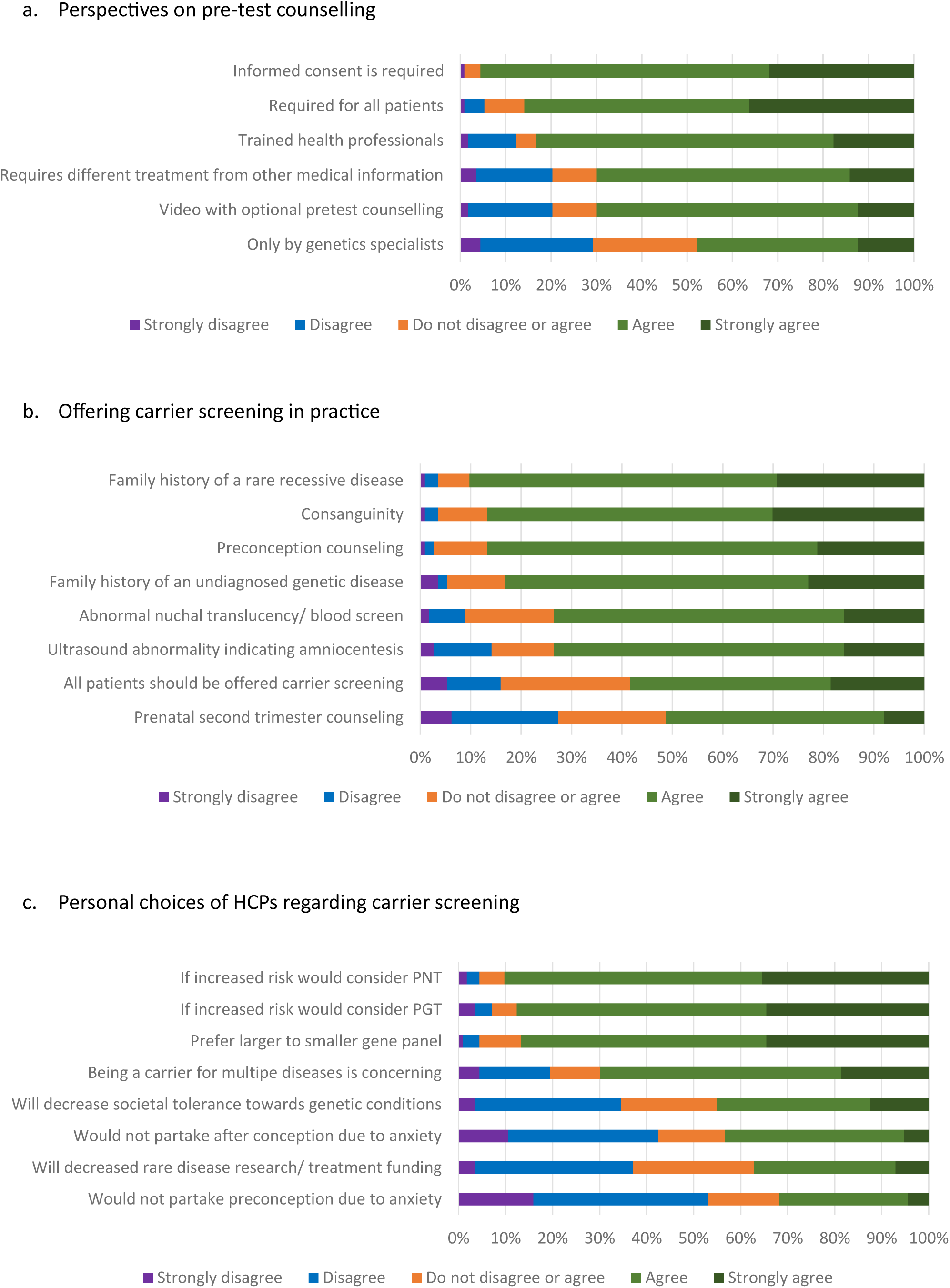
Healthcare professional perspectives towards carrier screening.

**Figure S4.**
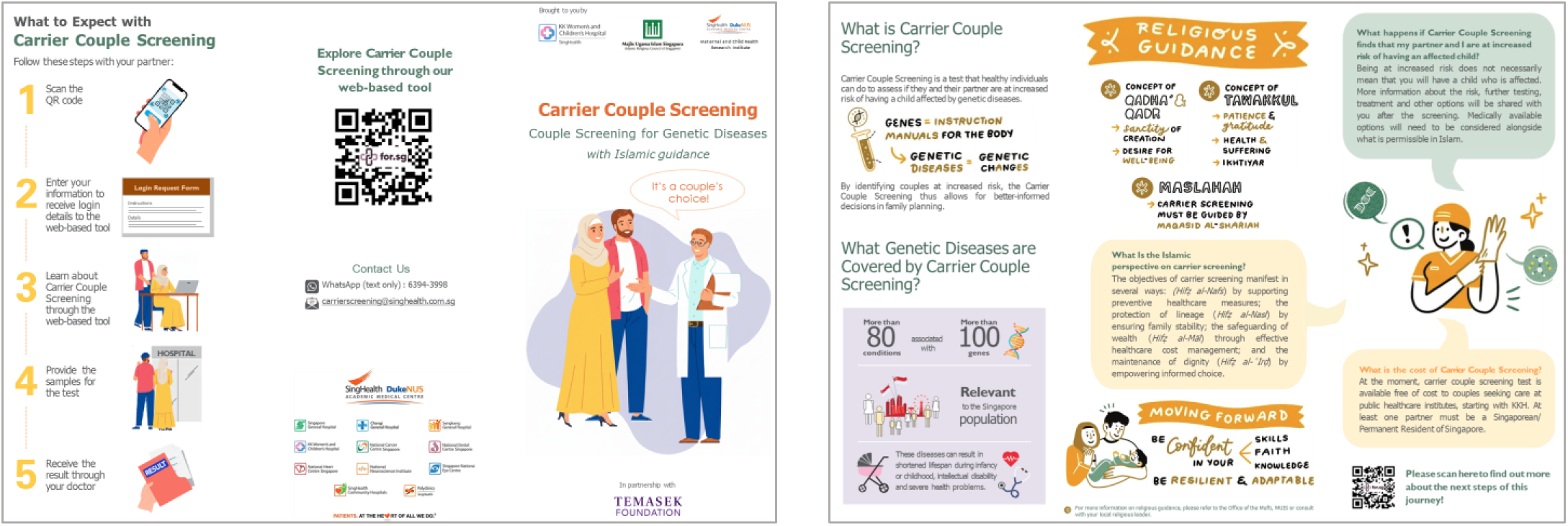
Modified Carrier Screening information brochure according to Islam guidance.

